# Longitudinal Analysis of Step Counts in Parkinson’s Disease Patients: Insights from a Web-Based Application

**DOI:** 10.1101/2023.11.22.23298898

**Authors:** Yishu Gong, Yuli Wang, Ziyang Wang, Xin Li, Yuan Gu

**Affiliations:** Harvard T.H. Chan School of Public Health, Harvard University, USA; Department of Biomedical Engineering, Johns Hopkins University, USA; Department of Computer Science, University of Oxford, UK; Department of Statistics, The George Washington University, USA

**Keywords:** Shiny, Web-based application, Parkinson’s disease, Longitudinal, Step count

## Abstract

Parkinson’s disease (PD) is a chronic neurological disorder that affects millions of people worldwide. One of the common motor symptoms associated with PD is gait impairment, leading to reduced step count and mobility. Monitoring and analyzing step count data can provide valuable insights into the progression of the disease and the effectiveness of various treatments. The generalized additive model (GAM) model presents the following variables: sex (Male vs. Female, p = 0.03), handedness (Right vs. Left/Both, p = 0.015), PD status of father (Yes vs. No, p = 0.056), COVID-19 status (Yes vs. No, p = 0.008), cohort (PD vs. healthy control, p < 0.0001), the cubic regression spline with three basis functions of age by cohorts (p<0.0001) and the random effect of the individual age trajectories (p = 0.0001) are statistically significant for daily step counts. A web application specifically tailored for step count analysis in PD patients was also developed and it provides a user-friendly interface for patients, caregivers, and healthcare professionals to track and analyze step count data, facilitating personalized treatment plans and enhancing the management of PD.

## 1 Introduction

Parkinson’s disease (PD) has emerged as the second most prevalent age-related neurodegenerative disorder worldwide, resulting in a substantial burden of disability and a significant increase in the risk of developing dementia and mortality. The overall prevalence rate of PD is 572 cases per 100,000 among individuals aged above 45 and over 93 cases per 100,000 among individuals aged 65 and older [1]. Despite significant progress in PD medications over the past half century, none can be classified as a cure. Although these drugs have improved in effectiveness and duration, they still do not fully alleviate PD symptoms and represent a long-term solution. Therefore, treatment options must be customized and tailored to meet the specific needs of each individual patient [2, 3, 4]. Additionally, the cause of PD remains unknown, and there are no established methods to prevent the disease. While several risk factors, such as pesticide exposure, have been identified, the confirmed causes of PD are genetic in nature. When PD is not linked to genetics, it is categorized as idiopathic (iPD), indicating that the precise cause is still unclear [5, 6].

Nowadays, the emergence of wearable solutions that provide low-cost, instant, convenient, and accurate measurements, such as smartwatches, fitness trackers, and health monitoring devices, have become increasingly popular due to their ability to seamlessly integrate into daily life. These wearables offer a wide range of functions, including tracking physical activity, monitoring vital signs, and even detecting certain health conditions. With their affordability, real-time data availability, and ease of use, wearable solutions have empowered individuals to take charge of their health and well-being in a more accessible and accurate manner [7].

Thanks to the reliability of wearable solutions, monitoring and preventing physical activity, especially during mild to moderate PD stages, holds particular significance. Previous research has extensively investigated clinically significant daily physical activities. They have provided evidence pertaining to physical performance metrics, such as gait, muscle strength, and step counts. Individuals with PD typically exhibit lower levels of physical activity compared to individuals of similar age without the condition. Even those who are newly diagnosed and haven’t started anti-Parkinson medications typically take around 5,000 steps per day if they are community-dwelling and able to walk independently. This falls short of the recommended 7,000 steps per day for older adults to maintain good health, highlighting the sedentary nature of the lifestyles often observed in people with PD [8]. Hence, step counts over time serve as a crucial indicator of motion in telemonitoring, with previous studies consistently demonstrating the continuous monitoring of step counts may serve as an important means of quantifying declining ambulatory behavior associated with disease progression or improved ambulatory behavior resulting from rehabilitation and medical and/or surgical interventions in persons with PD [9, 10]. These findings provide valuable insights for informing public health recommendations on physical activity, aiming to enhance prevention strategies for better overall health outcomes.

## 2 Related Work

Several studies have been conducted on PD patients, such as the WATCH-PD study, which was aimed to assess whether a combination of a smartwatch and smartphone could accurately measure early PD-related features [11], the Personalized Parkinson’s Project (PPP), a prospective single-center study of patients with early-stage PD in the Netherlands co-sponsored by Verily (Google Life Sciences) aiming to more precisely measure aspects of patient function and well-being [12]. Another landmark study is the Parkinson Progression Marker Initiative (PPMI) which is an ongoing longitudinal study conducted by The Michael J. Fox Foundation. It aims to identify biomarkers for PD progression, improving our understanding of the disease and enhancing the success of therapeutic trials targeting PD modification [13]. In 2018, Verily initiated a collaboration with PPMI to utilize Verily’s multi-sensor investigational wearable device. This device enables the continuous collection of data on movement, physiological measures, and environmental factors [14].

A lot of other studies, such as BETA-PD project [8], endeavor to delineate the physical activity (PA) profiles (three profiles: “Sedentary”, “Light Movers” and “Steady Movers”) of 301 individuals with PD. This pursuit aims to yield valuable insights for customizing PA interventions in a clinically meaningful manner. [15] aims to investigate the validity of various mHealth devices, including Google Fit, Health, STEPZ, Pacer, and Fitbit INC®, in estimating the step counts among a total of 34 individuals with idiopathic PD.

Despite the increasing interest and numerous studies dedicated to PD, there remains a paucity of studies investigating step count trends over an extended follow-up period and across various age groups among both healthy individuals and PD patients and large sample sizes in such research are lacking. Additionally, the absence of an open-source web application hinders clinicians’ ability to investigate the evolution of step count over time among different groups, such as variations based on factors like sex, COVID-19 status, hand dominance, and more. The lack of such a tool limits the exploration of potential correlations and insights that could contribute to personalized treatment approaches and further understanding of the disease.

Therefore, it is imperative to conduct a comprehensive longitudinal analysis of step counts, accounting for potential risk factors, using extensive and well-sourced long-term follow-up data, such as the PPMI study. On the other hand, although a couple of Shiny apps have been published online in our previous studies [16, 17], but they focus on different diseases, such as cancer or kidney failure, there is no accessible online platform for describing the baseline characteristics of PD patients or for modeling the time-varying trends in step counts between PD patients and healthy participants. This study addresses this gap by constructing a longitudinal model based on PPMI data and offering an online tool for clinical investigators to gain a more detailed visualization of these trends. Our online tool is published in the following link: https://baran-shad.shinyapps.io/RshinyPPMIstepcounts/.

## 3 Data and Methods

We requested access to the full breadth of individual-level PPMI longitudinal data from the Parkinson’s Progression Markers Initiative. The details of the PPMI study are available on the following link: https://www.ppmi-info.org/. A total of 353 subjects have recorded their step counts, with a follow-up period exceeding 2.11 years. The cumulative number of steps taken by each subject was recorded on an hourly basis. Our analysis includes additional demographic and characteristic factors such as sex, handedness (right or left/both), presence of Parkinson’s disease in parents (yes or no), and COVID-19 status, cohort (PD or healthy control). After incorporating these variables, excluding subjects who were not successfully enrolled or residing outside the United States (in a negative timezone) during the study, our final sample consists of 172 subjects.

### 3.1 Matching

After keeping as much as observations who have consistently step count records, we then employed a 1:4 (32 healthy control vs 126 PD) matching approach to extract subjects from the healthy control and PD groups, utilizing the “matchit” package in R. The matching model was specifically designed using the nearest neighbors approach, considering factors such as age, sex, handedness, and COVID-19 status to match the propensity scores of the two groups. The goal of the 1:4 matching approach was to pair each treated unit with the four closest control units, based on their respective propensity scores. By implementing this matching methodology, we aimed to establish comparability between the treated and control groups, enhancing the validity of our analysis and mitigating potential confounding factors. After merging with the number of presence of Parkinson’s disease in parents, the final analysis data set contains 32 healthy control and 126 PD patients.

### 3.2 Time Binning

To account for the varying decimal values of ages represented in days, we employed the binning method in our study. This approach facilitated the creation of equally spaced time bins, specifically using intervals of 3 months (90 days). For instance, age groups ranging from 66 to 67 were binned into intervals of 0.25. This resulted in age bin labels such as 66.25, 66.50, 66.75, and 67. By grouping the data in this manner, we were able to increase the sample size within each binned age interval and aggregate step counts within each age bin. By utilizing binning in conjunction with cumulative step counts, we aimed to capture and analyze the overall activity patterns across different age intervals in a more manageable and informative manner. This approach facilitated the examination of age-related patterns and trends with increased resolution, providing valuable insights into the relationship between age and other variables under investigation.

### 3.3 Baseline Characteristics and descriptive plots

There are a total of 32 healthy control subjects and 126 Parkinson’s patients (Table 1). Among those subjects, there are 61 females and 97 males. In previous multiple studies [18, 19], left-handedness is also considered a potential risk factor for PD, we incorporated handedness as a potential factor in our analysis. Within our data set, we have a total of 131 right-handed individuals, and 27 left-handed individuals or who exhibit ambidextrous tendencies. The study coincided with the outbreak of the pandemic, and it’s important to note that COVID-19 status poses significant risks, particularly for the elderly population, including those individuals who are already diagnosed with Parkinson’s disease. COVID-19 status is also collected and there are a total of 30 COVID-19 patients. Another unconventional risk factor under consideration is the PD status of both the father and the mother. A total of 17 patients have fathers with Parkinson’s disease, and 11 patients have mothers with Parkinson’s disease.

**Table 1.** Baseline characteristics by healthy control and PD groups.

|  | Healthy Control (n=32) | Parkinson's Disease (n=126) | P-value |
| --- | --- | --- | --- |
| COVID-19 status |  |  |  |
| No | 23 (72%) | 105 (83%) | 0.14 |
| Yes | 9 (28%) | 21 (17%) |  |
| Sex |  |  |  |
| Female | 12 (37%) | 49 (39%) | 0.8854 |
| Male | 20 (63%) | 77 (61%) |  |
| Handedness |  |  |  |
| Right | 24 (75%) | 107 (85%) | 0.1831 |
| Left/Both | 8 (25%) | 19 (15%) |  |
| PD status of father |  |  |  |
| No | 32 (100%) | 109 (87%) | 0.0278 |
| Yes | 0 (0%) | 17 (13%) |  |
| PD status of mother |  |  |  |
| No | 32 (100%) | 115 (91%) | 0.0831 |
| Yes | 0 (0%) | 11 (9%) |  |

**Table 2.** Comparison of Variables in GAM and LME Models.

| GAM |  |  | LME |  |  |
| --- | --- | --- | --- | --- | --- |
| Variable | Coefficient | p-value | Variable | Coefficient | p-value |
| Sex | 388.8 | 0.030 | Sex | -19.7 | 0.918 |
| Handedness<br>(Yes:Right; No:Left/Both) | 564.5 | 0.015 | Handedness<br>(Yes:Right; No:Left/Both) | 853.0 | < 0.001 |
| PD status of father | 588.4 | 0.056 | PD status of father | 1059.7 | 0.002 |
| PD status of mother | -251.9 | 0.471 | PD status of mother | -884.8 | 0.010 |
| COVID-19 | 538.5 | 0.008 | COVID-19 | 768 | < 0.001 |
| Cohort<br>(PD: 1; Healthy Control:2) | 1906.6 | < 0.0001 | Cohort<br>(PD: 1; Healthy Control:2) | 3234.9 | < 0.0001 |
| Smoothing on age | - | < 0.0001 | Random effect of age | - | < 0.0001 |
| Random effect of smoothing trajectory on age | - | 0.0001 |  |  |  |
| AIC | - | 16465.8 | AIC | - | 16513.28 |

The p-values for the chi-square test are also presented in Table 1, to present whether there exists a statistically significant difference between the two groups (PD and healthy control) across different variables. Notably, most of these p-values are greater than 0.05, indicating a lack of statistical significance in difference. The variable “PD status of Father” boasts a p-value of 0.0278, indicating a significant difference in the PD status of fathers between the healthy control and PD groups. For other baseline characteristics, all the p-values > 0.05, indicating there is no significant difference in those characteristics between PD and healthy control groups and we may assume that the variables are evenly distributed between the two groups.

In Figure 1, the step counts over time are depicted for each subject, with the left side representing the healthy control group and the right side illustrating the PD group. Moving on to Figure 2, it showcases the change in step counts from the baseline across time for each subject. The left side pertains to the healthy control group, while the right side corresponds to the PD group. When comparing the two groups in Figure 1, it becomes evident that individuals in the healthy control group take more daily steps in contrast to PD patients. Furthermore, in Figure 2, we observe greater variability among PD patients over time as compared to the healthy control group. This suggests the possibility of a significant decline in daily step counts as PD progresses.

**Fig. 1.**
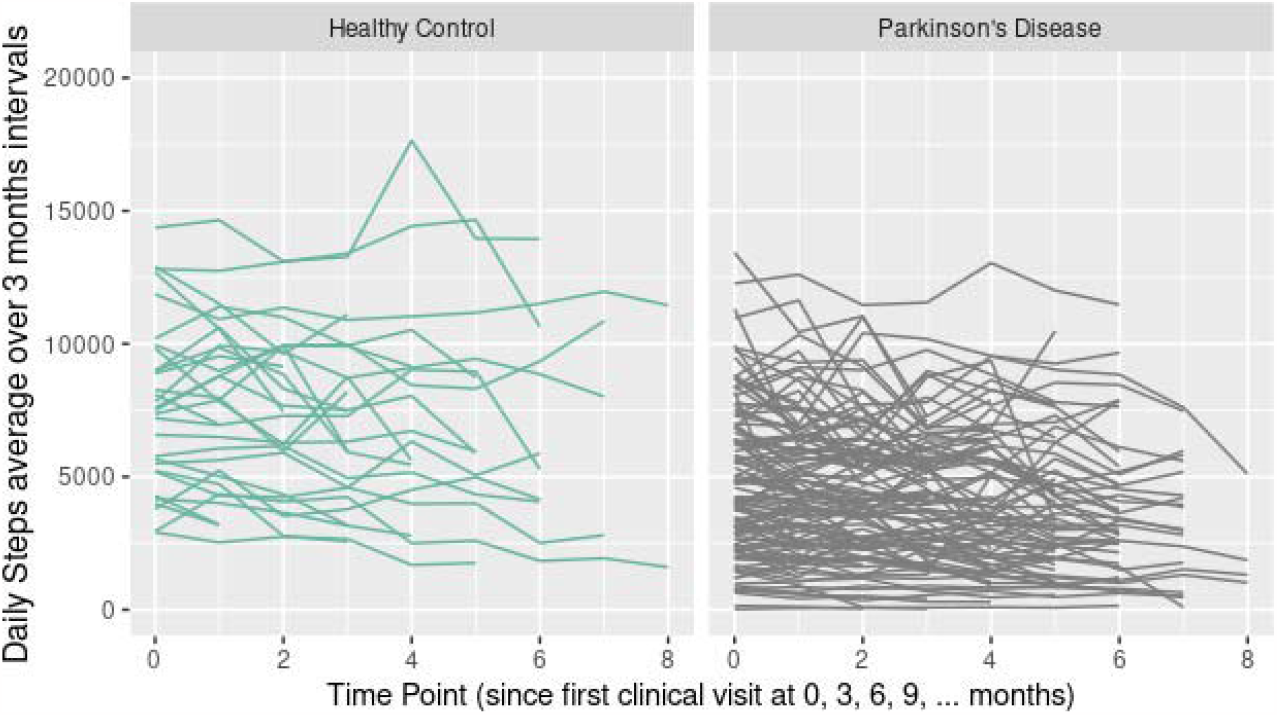
Trajectories of daily steps over time.

**Fig. 2.**
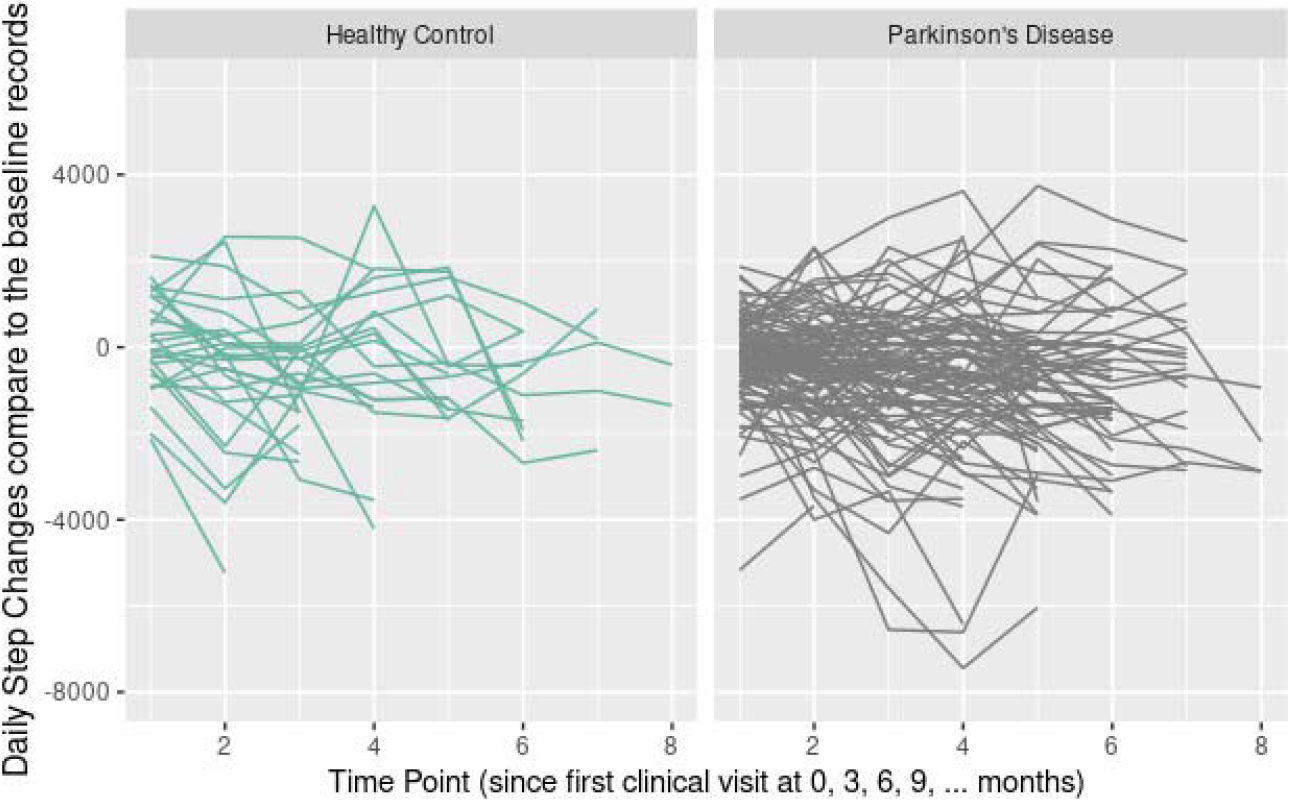
Trajectories of daily steps change compared to the baseline records over time.

Figure 3 shows a comparison between the PD and the healthy control groups using boxplots over time. The time variables are using a 3-month time interval unit. A more precise statistical representation can be observed through the box plots, which not only display the mean and range but also highlight any outliers. Through these box plots, the discernible distinction in step counts between the two groups becomes evident at every time point. This trend of higher step counts among individuals in the healthy control group as opposed to the PD group is consistently depicted in the three figures. Further descriptive plots are available within our application app for more comprehensive insights.

**Fig. 3.**
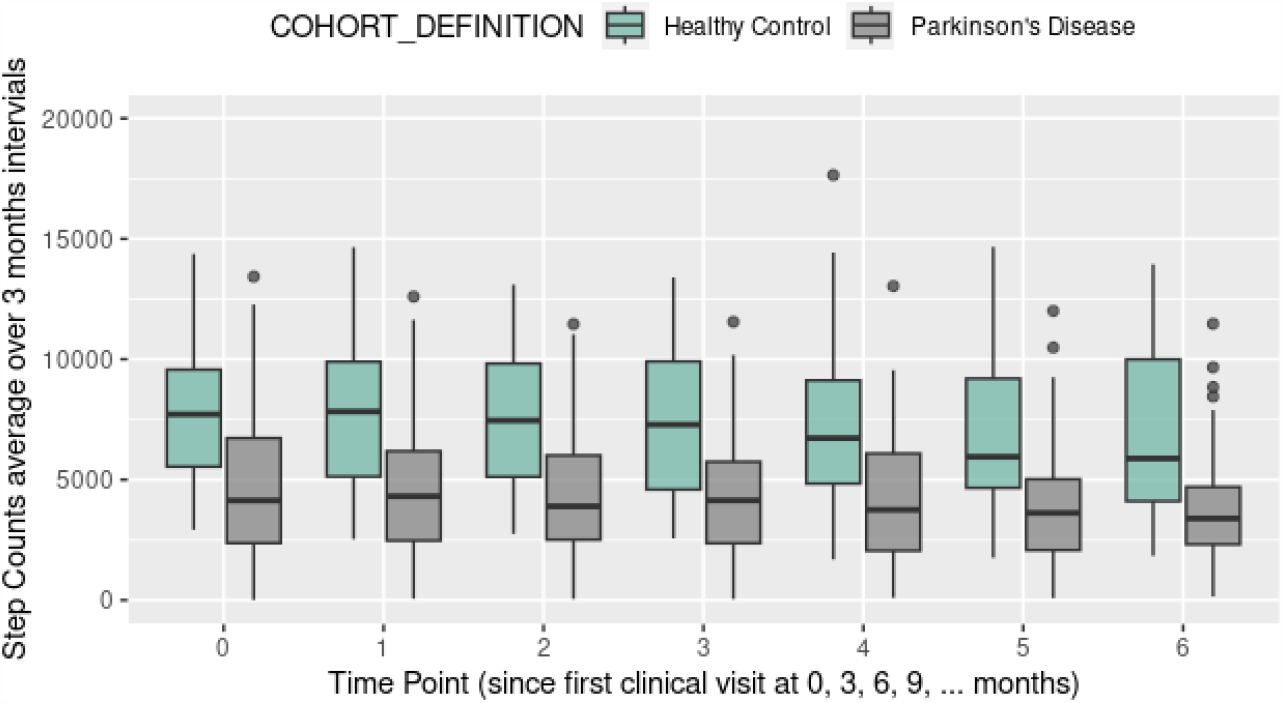
Box plots of daily steps average over 3 months by different time (3 months intervals).

## 4 Generalized Additive Model Results

The standard linear mixed-effect model (LME) is a common choice for longitudinal data analysis. However, it may not be able to effectively capture the curvature of time-varying trajectories, particularly when quadratic relationships exist in the repeated measurements. To address this limitation, we have employed the generalized additive model (GAM) in our study. Unlike the standard linear model that relies solely on linear feature contributions, GAM is constructed as a sum of smooth functions of features. This flexibility allows GAM to better accommodate and model complex, non-linear relationships within the data, enhancing the accuracy and interpretability of our analysis.

In the GAM model, we used daily step counts averages over 7 day period (with at least 6 days) to accormmodate for weekly variability. The daily step counts (range is 6 - 17645 /per day) are used as the dependent variables and the independent variables are sex, handedness, PD status of father, PD status of mother, COVID-19 status, cohort: PD vs healthy Control, also we used the cubic regression spline with three basis functions of age by cohorts and also the random effect of the individual age trajectories.

The result comparison between GAM and LME model is displayed in 2. We observe that several variables including sex (p-value = 0.03 in GAM, 0.918 in LME), handedness (p-value = 0.015 in GAM, < 0.001 in LME), PD status of the father (p-value = 0.056, close to the significant level in GAM, 0.002 in LME), COVID-19 status (p-value = 0.008 in GAM, < 0.001 in LME), cohort (p-value < 0.0001 in GAM, < 0.0001 in LME), as well as the smoothing curve on age (p-value < 0.0001), and the random effect of the smoothing trajectory on age (p-value = 0.0001), all demonstrate statistical significance or close to significant in relation to step counts except PD status of mother with a p-value of 0.47 in GAM but 0.01 in LME. The Akaike information criterion (AIC) metric to compare the two models is also presented. The AIC of GAM is lower than that of LME, indicating a slightly better fit in the GAM. Althogh slight difference, both models revealed similar and significant predictors, including handedness, family history of PD, COVID-19 status, cohort, as well as the smoothing or random effects of age. Notably, among these features, handedness displays a positive coefficient, implying that right-handed individuals exhibit higher step counts when compared to their left-handed counterparts or those who are ambidextrous. And as the study extended beyond the COVID-19 period, it’s noteworthy that COVID-19 status is another factor that statistically affects daily step counts.

Figure 4 below depicts the step counts estimated by our GAM model. These results are presented as smooth curves, with green curves representing the healthy control group and gray curves for the PD group. Additionally, the original daily step data is overlaid as connected lines, providing background information to enhance the clarity and comprehensibility of the visualization. This visualization reinforces the observation that individuals within the PD group exhibit significantly fewer step counts in comparison to the control group, also in both groups, the step counts are decreasing over age, especially after age 70. The fitted linear lines from the LME are also presented in Figure 5, when compared to the fitted curve from the GAM, these linear lines exhibit a similar overall trend in step counts over age in both groups. However, the LME does not capture the curvature in the trend. Morefigures are displayed within the Shiny application, allowing clinicians to customize the way the figures are presented by making selections from a drop-down menu.

**Fig. 4.**
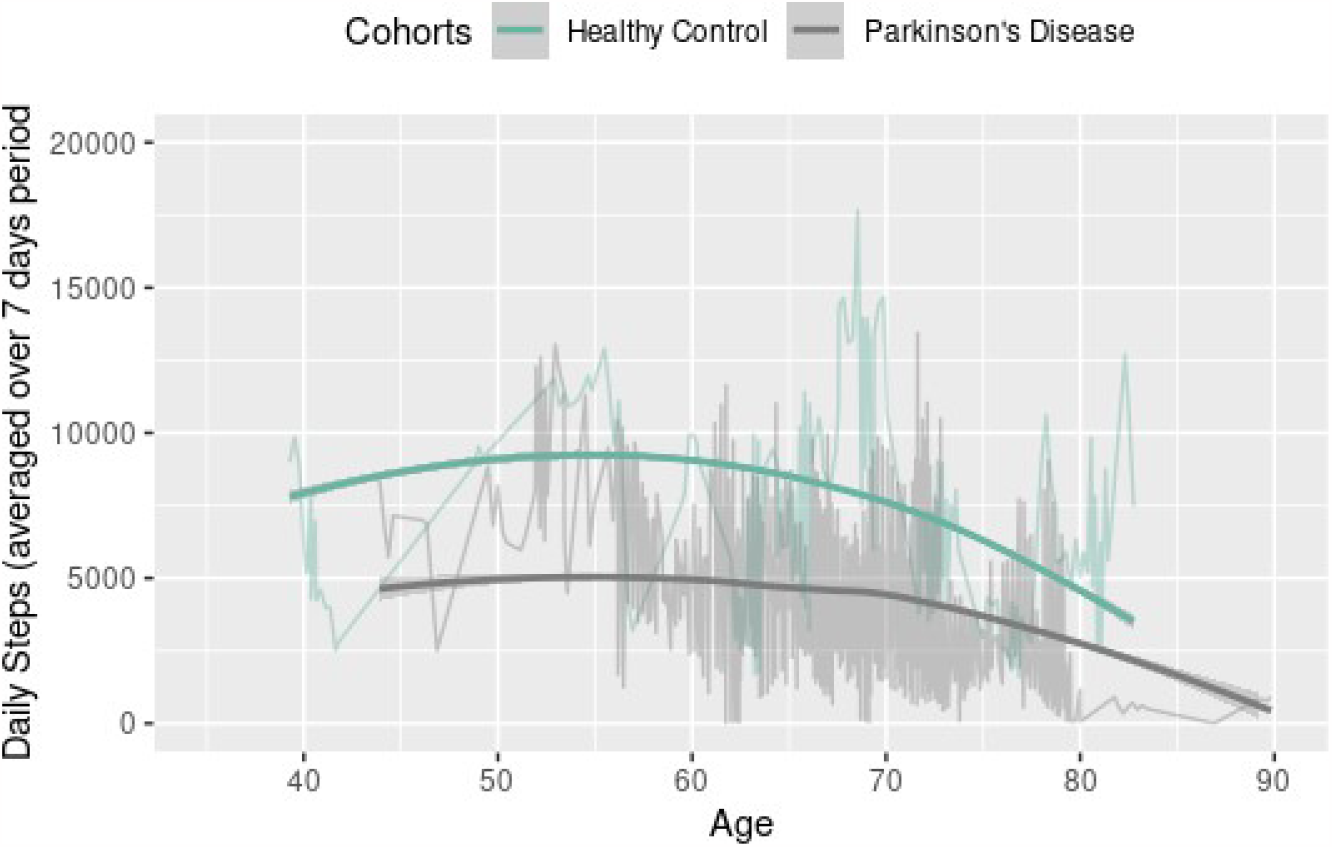
Daily Steps comparison between healthy control (Blue) and PD groups over Age Curves: GAM fitted curve; Connected lines: Original Daily Steps.

**Fig. 5.**
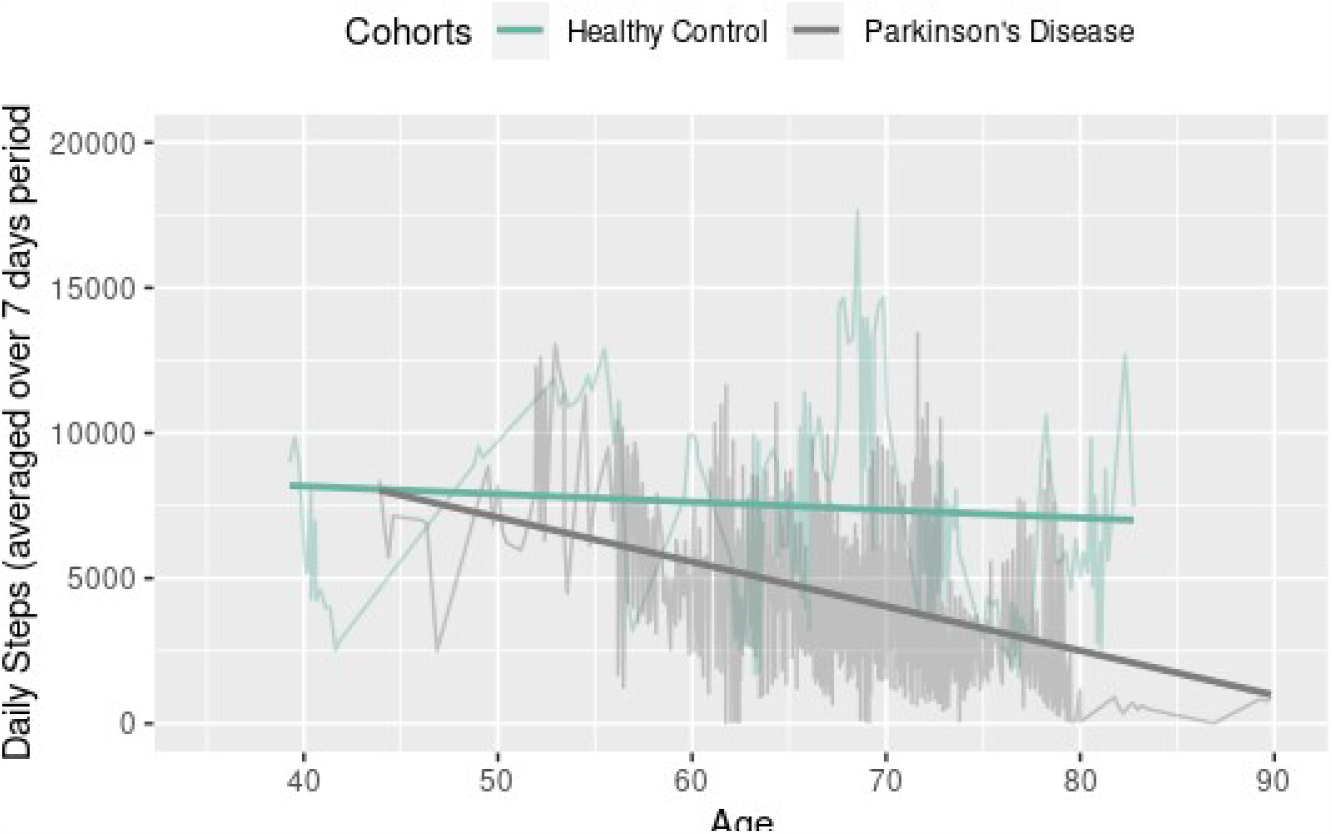
Daily Steps comparison between healthy control (Blue) and PD groups over Age Curves: LLM fitted line; Connected lines: Original Daily Steps.

We conducted additional investigations into the results of the GAM by examining different cohort groups, as summarized in Table 3. It’s important to note that in the descriptive statistics presented in Table 1, there were no instances of PD family history among the healthy control group. Hence, we excluded the two variables related to PD status of father and mother from the model for the healthy control group. Additionally, it’s worth mentioning that in the healthy control group, there is a notably higher proportion of individuals identified as “Left/Both” (25%) in comparison to the PD group. In the case of PD patients, it’s notable that handedness does not appear to be a significant factor. This observation could be attributed to the smaller proportion of individuals in the “left/both” group, which accounts for only 15% (n=19) out of the 126 PD subjects. However, the effects of the other variables remain consistent with those observed in the GAM for all participants. For the control participants, it’s important to highlight that only sex does not exhibit a significant effect on step counts over time. In contrast, handedness, COVID-19 status, the age smoothing term, and its random effect all demonstrate significant effects on step counts over time in this group.

**Table 3.**
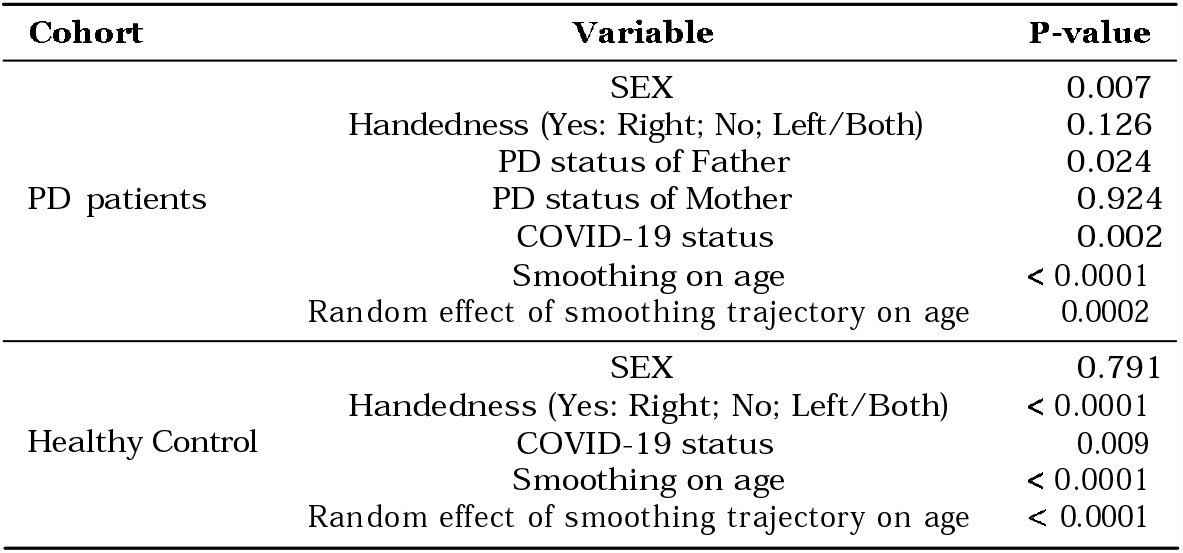
GAM for different cohort groups.

| Cohort | Variable | P-value |
| --- | --- | --- |
| PD patients | SEX | 0.007 |
|  | Handedness (Yes: Right; No: Left/Both) | 0.126 |
|  | PD status of Father | 0.024 |
|  | PD status of Mother | 0.924 |
|  | COVID-19 status | 0.002 |
|  | Smoothing on age | < 0.0001 |
|  | Random effect of smoothing trajectory on age | 0.0002 |
| Healthy Control | SEX | 0.791 |
|  | Handedness (Yes: Right; No: Left/Both) | < 0.0001 |
|  | COVID-19 status | 0.009 |
|  | Smoothing on age | < 0.0001 |
|  | Random effect of smoothing trajectory on age | < 0.0001 |

## 5 Conclusion

Although numerous studies have concentrated on PD, there are only a limited number of investigations that have delved into the step counts of PD patients over a long time period. This scarcity is largely attributed to the substantial volume of daily data records involved, making continuous follow-up challenging to execute effectively. Even in the most comprehensive PPMI study, the number of participants in the healthy control group followed up for up to 2 years remains limited compared to the PD groups. This limitation leads to unbalanced data between the two groups and may restrict our model’s ability to provide a more generalized outcome. Our analysis adeptly managed the voluminous and intricate daily step count data obtained from PPMI.

Throughout our investigation, we pinpointed several statistically significant factors that impact step counts. These factors include gender, COVID-19 status, handedness, the family history of PD status, as well as the application of cubic spline regression on age and the incorporation of individual age as a random effect. Our GAM model analysis revealed that when adjusting the effects of other variables, handedness bears a positive coefficient which suggests that individuals who are left-handed or ambidextrous tend to record lower step counts in comparison to their right-handed counterparts.

This discovery aligns with previous studies and also serves as a catalyst for our future research into imaging disparities between right-handed and left-handed individuals. The potential analysis methods to be employed include assessing different time courses of functional loci between right-handed and left-handed participants through image segmentation via location network combining with 3D U-net or utilizing augmented pyramid networks on an encoder-decoder multi-class segmentation framework such as QAP-NET, RAR-U-Net, etc. [20, 21, 22, 23, 24, 25]. Gender is another associated factor, with men showing higher daily step counts compared to women. The curvature of cubic regression over age is also a significant factor; as age increases, daily step counts tend to decrease, especially after reaching the age of 70. Additionally, COVID-19 status is a statistically significant factor that influences daily step counts. Another intriguing discovery pertains to the family history of PD. The PD status of the father exhibits a p-value of 0.056, indicating its proximity to the significance threshold. Due to the limitation of the data, there is only a relatively small subset of patients whose parents also have PD, thereby limiting the comprehensive examination of familial influences, further relevant data needed to be collected to conduct a more comprehensive investigation.

In addition to the aforementioned findings, we developed an R Shiny web application that facilitates investigators in iteratively exploring various descriptive plots. This application enables the visualization of step count trajectories over time for different groups, providing a dynamic tool for enhanced data analysis and interpretation.

However, besides the above limitations we have mentioned, the healthy control group’s sample size is relatively small in this study, potentially constraining the robustness of comparative analyses. Additionally, the number of independent variables considered in our study is limited, which might not encapsulate the entirety of potential contributing factors. To address these limitations and achieve a more comprehensive understanding, it becomes evident that a larger and more diverse dataset, coupled with an extended follow-up period, is necessary. This expansion of data and temporal coverage will not only provide a broader context for analysis but also offer the opportunity to explore the nuances of PD progression and its interplay with various variables in greater depth.

## Data Availability

All data produced are available online at

https://www.ppmi-info.org/

## 7 Data Acknowledgement

We like to thank all patients and healthy volunteers for participating in the PPMI study, all investigators who contributed, and the Michael J. Fox Foundation. Data used in the preparation of this article were obtained [on May, 23, 2023] from the Parkinson’s Progression Markers Initiative (PPMI) database (www.ppmi-info.org/access-data-specimens/download-data), RRID:SCR 006431. For up-to-date information on the study, visit www.ppmi-info.org

## References

[1] Marras, C. et al. Prevalence of parkinson’s disease across north america. NPJ Parkinson’s disease 4, 21 (2018).

[2] Sioka, C., Fotopoulos, A. & Kyritsis, A. P. Recent advances in pet imaging for evaluation of parkinson’s disease. European journal of nuclear medicine and molecular imaging 37, 1594–1603 (2010).

[3] Lajurkar, S. V., Nade, V. S., Bhoir, P. V. & Kardus, N. A. Current status and novel strategies for prevention and treatment of parkinson’s disease (2018).

[4] Stoker, T. B. & Barker, R. A. Recent developments in the treatment of parkinson’s disease. F1000Research 9 (2020).

[5] Mahlknecht, P., Marini, K., Werkmann, M., Poewe, W. & Seppi, K. Prodromal parkinson’s disease: hype or hope for disease-modification trials? Translational neurodegeneration 11, 1–13 (2022).

[6] Puigrós, M. et al. Cell-free mitochondrial dna deletions in idiopathic, but not lrrk2, parkinson’s disease. Neurobiology of Disease 174, 105885 (2022).

[7] Channa, A., Popescu, N. & Ciobanu, V. Wearable solutions for patients with parkinson’s disease and neurocognitive disorder: a systematic review. Sensors 20, 2713 (2020).

[8] von Rosen, P., Hagströmer, M., Franzɾn, E. & Leavy, B. Physical activity profiles in parkinson’s disease. BMC neurology 21, 1–8 (2021).

[9] Cavanaugh, J. T. et al. Capturing ambulatory activity decline in parkinson disease. Journal of neurologic physical therapy: JNPT 36, 51 (2012).

[10] Master, H. et al. Association of step counts over time with the risk of chronic disease in the all of us research program. Nature medicine 28, 2301–2308 (2022).

[11] Adams, J. L. et al. Using a smartwatch and smartphone to assess early parkinson’s disease in the watch-pd study. npj Parkinson’s Disease 9, 64 (2023).

[12] Diao, J. A., Raza, M. M., Venkatesh, K. P. & Kvedar, J. C. Watching parkinson’s disease with wrist-based sensors. NPJ Digital Medicine 5, 73 (2022).

[13] Marek, K. et al. The parkinson progression marker initiative (ppmi). Progress in neurobiology 95, 629–635 (2011).

[14] Atri, R. et al. Deep learning for daily monitoring of parkinson’s disease outside the clinic using wearable sensors. Sensors 22, 6831 (2022).

[15] de Carvalho Lana, R., de Paula, A. R., Silva, A. F. S., Costa, P. H. V. & Polese, J. C. Validity of mhealth devices for counting steps in individuals with parkinson’s disease. Journal of Bodywork and Movement Therapies 28, 496–501 (2021).

[16] Gu, Y. et al. Enhancing kidney failure analysis: Web application development for longitudinal trajectory clustering. medRxiv 2023–05 (2023).

[17] Gu, Y. et al. Unveiling breast cancer risk profiles: A comprehensive survival clustering analysis empowered by an interactive online tool for personalized medicine. medRxiv 2023–05 (2023).

[18] Yust-Katz, S., Tesler, D., Treves, T. A., Melamed, E. & Djaldetti, R. Handedness as a predictor of side of onset of parkinson’s disease. Parkinsonism & related disorders 14, 633–635 (2008).

[19] van der Hoorn, A., Burger, H., Leenders, K. L. & de Jong, B. M. Handedness correlates with the dominant parkinson side: A systematic review and meta-analysis. Movement Disorders 27, 206–210 (2012).

[20] Wang, Y. et al. Automated ventricle parcellation and evan’s ratio computation in∽ pre-∽ and∽ post-surgical∽ ventriculomegaly. arXiv preprint arXiv:2303.01922 (2023).

[21] Wang, Y. & Yi, J. Deep learning-based image registration method: with application to scanning laser ophthalmoscopy (slo) longitudinal images. In Medical Imaging 2023: Image Processing, vol. 12464, 601–605 (SPIE, 2023).

[22] Wang, Z. & Voiculescu, I. Quadruple augmented pyramid network for multi-class covid-19 segmentation via ct. In 2021 43rd Annual International Conference of the IEEE Engineering in Medicine & Biology Society (EMBC), 2956–2959 (IEEE, 2021).

[23] Wang, Z., Zhang, Z. & Voiculescu, I. Rar-u-net: a residual encoder to attention decoder by residual connections framework for spine segmentation under noisy labels. In 2021 IEEE International Conference on Image Processing (ICIP), 21–25 (IEEE, 2021).

[24] Song, J., Gu, Y. & Kumar, E. Chest disease image classification based on spectral clustering algorithm. Research Reports on Computer Science 77–90 (2023).

[25] Zhou, P. et al. Automatically detecting bregma and lambda points in rodent skull anatomy images. PloS one 15, e0244378 (2020).

